# Remote digital measurement of visual and auditory markers of Major Depressive Disorder severity and treatment response

**DOI:** 10.1101/2020.08.24.20178004

**Authors:** Isaac R. Galatzer-Levy, Anzar Abbas, Vijay Yadav, Vidya Koesmahargyo, Allison Aghjayan, Serena Marecki, Miriam Evans, Colin Sauder

## Abstract

**Objectives:** Multiple machine learning-based visual and auditory digital markers have demonstrated associations between Major Depressive Disorder (MDD) status and severity. The current study examines if such measurements can quantify response to antidepressant treatment (ADT) with selective serotonin reuptake inhibitors (SSRIs) and serotonin-norepinephrine uptake inhibitors (SNRIs).

**Methods:** Visual and auditory markers were acquired through an automated smartphone task that measures facial, vocal, and head movement characteristics across four weeks of treatment (with timepoints at baseline, 2 weeks, and 4 weeks) on ADT *(n* = 12). The Montgomery-Asberg Depression Rating Scale (MADRS) was collected concordantly through clinical interviews to confirm diagnosis and assess changes in MDD severity.

**Results:** Patient responses to ADT demonstrated clinically and statistically significant changes in the MADRS F(2,34) = 51.62, *p* <.0001. Additionally, patients demonstrated significant increases in multiple digital markers including facial expressivity, head movement, and amount of speech. Finally, patients demonstrated significant decreased frequency of fear and anger facial expressions.

**Conclusion:** Digital markers associated with MDD demonstrate validity as measures of treatment response.

## 1 Introduction

Patients with Major Depressive Disorder (MDD) are both heterogeneous in their clinical presentation and their response to antidepressant treatments (ADTs) (Lux and Kendler, 2010; Ostergaard et al., 2011). It is theorized that treatment effects may be obfuscated because MDD measurements combine heterogeneous symptoms that reflect distinct neurobiological and social processes while pharmacological treatments target specific neurobiological processes such as serotonergic tone. For example, patients with different subtypes of MDD, such as cognitive and neurovegetative phenotypes, have demonstrated differential treatment response to distinct classes of ADTs. (Jaracz et al., 2015; Uher et al., 2009) As such, there are significant efforts to refocus treatment research on measures that match the underlying neurobiological treatment target (Cuthbert and Insel, 2013). Disentangling the heterogeneity in MDD can lead to better risk and treatment response assessment by shifting the focus of investigation to narrow phenotypes that reflect the underlying neurological deficit and target of treatment (Cuthbert and Insel, 2013; Insel, 2014).

The use of digital measurements that relate to underlying biological phenotypes, termed digital phenotyping (Insel, 2017), has been proposed as a methodology to improve measurement of underlying illness by capturing digital proxy measures of clinical functioning. An example of digital phenotyping is the measurement of activity as a proxy measure of mood or anxiety states using actigraphy or geolocation captured from an individual’s smartphone (Onnela and Rauch, 2016; Torous et al., 2017). While novel measurements are promising, validation is required before such metrics can be interpreted clinically. Key steps to validation include comparison to traditional clinical measures, both cross-sectionally and as they change with the disease or treatment course (Coravos et al., 2019). Such measures should strive for ease of collection and increased sensitivity to facilitate frequent, accurate assessment and should be validated in relationship to narrower biological phenotypes and treatment targets than those that traditional endpoints assess. This will ultimately lead to improved, dynamic treatment research and clinical decision making (Torous et al., 2017) based on modulation of underlying neurobiological deficits (Lenze et al., 2020).

Based on prior knowledge, visual and auditory data sources represent a compelling direction for objective measurement of patient functioning in MDD. Beginning with observations by Emil Kraepelin, patients with depression have been shown to produce slowed and spaced out speech, where they appear to "become mute in the middle of a sentence" and demonstrate altered facial behavior, regarding which he states, “the facial expression and the general attitude are sleepy and languid” (Kraepelin, 1907). These clinical observations by Kraepelin have been corroborated and extended with standardized methods to assess facial expressions, vocal characteristics, and movement patterns using audio and video data sources. The same paucity of speech has been observed in acutely suicidal patients (Cummins et al., 2015). Indeed, both speech and facial/bodily movement represent sensitive biological outputs that change with physiological and cognitive variability (Cummins et al., 2015; Dagum, 2018; Jabbi and Keysers, 2008).

A number of visual and auditory characteristics that correspond to known MDD symptoms can now be directly quantified. This includes reduced gross motor activity (Buyukdura et al., 2011), slumped posture (Kim et al., 2018), reduced head movement variability (Alghowinem et al., 2013; Dibeklioglu et al., 2018; Kim et al., 2018), reduced facial expressivity (Girard et al., 2013), reduced speech production (Garcia-Toro et al., 2000), and increased negative affect (Berenbaum and Oltmanns, 2005; Ekman and Rosenberg, 2005). The automated measurement of these clinical features introduces the possibility of objective automated assessment. Given that audio and video data sources can be captured remotely, this further introduces the possibility of greatly scaling the reach and frequency of assessment. Increased scale and objectivity can facilitate increased accuracy and accessibility of clinical risk and treatment response assessments.

Serotonin signaling deficits represent a primary biological target for treatment in MDD. Serotonergic tone mechanistically impacts motor functioning directly through interactions with dopamine and norepinephrine signalling (El Mansari et al., 2010; Herrera-Guzmán et al., 2009; Morrissette and Stahl, 2014). Postmortem comparison of suicides compared to controls demonstrates significant reductions of brain serotonin (Stockmeier, 2003; Stockmeier et al., 1998). More specific mapping of mRNA expression patterns demonstrates reduced expression of Serotonin mRNA subtypes that are relatively widespread and other subtypes that are specific to the frontopolar cortex amygdala circuitry (Anisman et al., 2008). This circuitry governs the expression and regulation of threat and anxiety (Gold et al., 2015).

In the current study, we tested for changes in digital markers of (1) facial expressivity, (2) negative affect measured in facial expression, (3) pauses in speech, and (4) head movement activity. We hypothesized significant positive change in measures of negative affect and significant negative change in measures of motor functioning (such as percentage of audio with detected speech, facial expressivity, and head movement activity) in patients diagnosed with MDD over the first 4 weeks of ADT.

## 2. Methods

### 2.1 Study Participants

Participants were identified through advertisements posted on social media. Individuals who self-identified as experiencing depression were screened over the telephone to assess depression symptoms. Potentially eligible subjects were then scheduled for an in-person pre-screening visit with a clinician to assess primary eligibility criteria. Individuals who met criteria consented to participate in a screening assessment with a psychological rater, which included the Mini-International Neuropsychiatric Interview (MINI), Structured Interview Guide for the Montgomery-Asberg Depression Rating Scale (SIGMA-MADRS), Columbia Suicide Severity Rating Scale (C-SSRS), and the Quick Inventory of Depressive Symptomatology Self Report (QIDS-SR16).

To be included in the study, subjects had to meet DSM-5 criteria for single or recurrent MDD based on the MINI with a current major depressive episode of ≥ 8 weeks and a MADRS total score of ≥ 20. Participants must have also been, in the opinion of the study psychiatrist, medically stable and a good candidate for treatment with a monoamine ADT. Key exclusion criteria included significant medical complications (e.g., uncontrolled cardiac or endocrine disorders, diagnosis or treatment for cancer within the past 2 years), significant psychiatric complications (e.g., suicidal behavior that in the judgment of the investigator confers significant risk, other primary psychiatric diagnosis, substance use disorder), or the use of certain prohibited concomitant medications (e.g., prescription painkillers [opioids]).

Participants who met screening eligibility criteria subsequently completed a visit with a study psychiatrist and were prescribed an ADT consistent with standard of care. Participants who demonstrated significant decreases in depression severity, indicated by a 30% reduction in MADRS total score over 4 weeks of ADT, were included in the sample (n = 18). The sample included 7 men and 11 women (mean age = 30.2 ± 8.6), who met criteria for MDD. The mean BMI was 28.7 ± 5.6. Baseline total MADRS scores ranged from 25 to 45 (mean = 34.1 ± 4.9).

### 2.2 Treatment and Assessment Conditions

All patients were prescribed either an SSRI or SNRI at variable doses based on the clinician’s discretion. Treatment response was measured at bi-weekly intervals using two independent assessments described below.

### 2.3 Assessments

#### 2.3.1 Clinical Assessment

Montgomery-Asberg Depression Rating Scale (MADRS): The MADRS is a 10-item clinician administered scale for the measurement of MDD with validated clinical cut points for Severe (>34), Moderate (17-34), Mild (7-19), and Asymptomatic (< 7) depression. The MADRS has demonstrated validity as a sensitive measure of ADT response (Montgomery et al., 1985). The MADRS was administered by trained psychological raters with prior scale experience at Week 0 (Baseline) and at approximately 2 and 4 weeks post treatment initiation.

#### 2.3.2 Remote smartphone-based video assessments

All participants were asked to download the AiCure app (AiCure, LLC, New York, NY www.aicure.com) on their personal smartphone for measurement of digital markers of MDD. They were then trained by the study team on how to use the app to participate in remote assessments.

Participants completed remote assessments once a week for the length of the study. The assessment consisted of a smartphone-based adaptation of a paradigm to examine emotional valence in response to varied emotional imagery (Carretié et al., 2019; Stockmeier, 2003; Stockmeier et al., 1998). Specifically, at each assessment time point they were prompted to view images selected at random from the Open Affective Standardized Image Set (OASIS) from one of three emotional valence categories: positive valence, neutral valence, and negative valence. Participants were asked to observe each image and then describe what they saw and how it made them feel for a minimum of 10 seconds (Figure 1).

**Figure 1:**
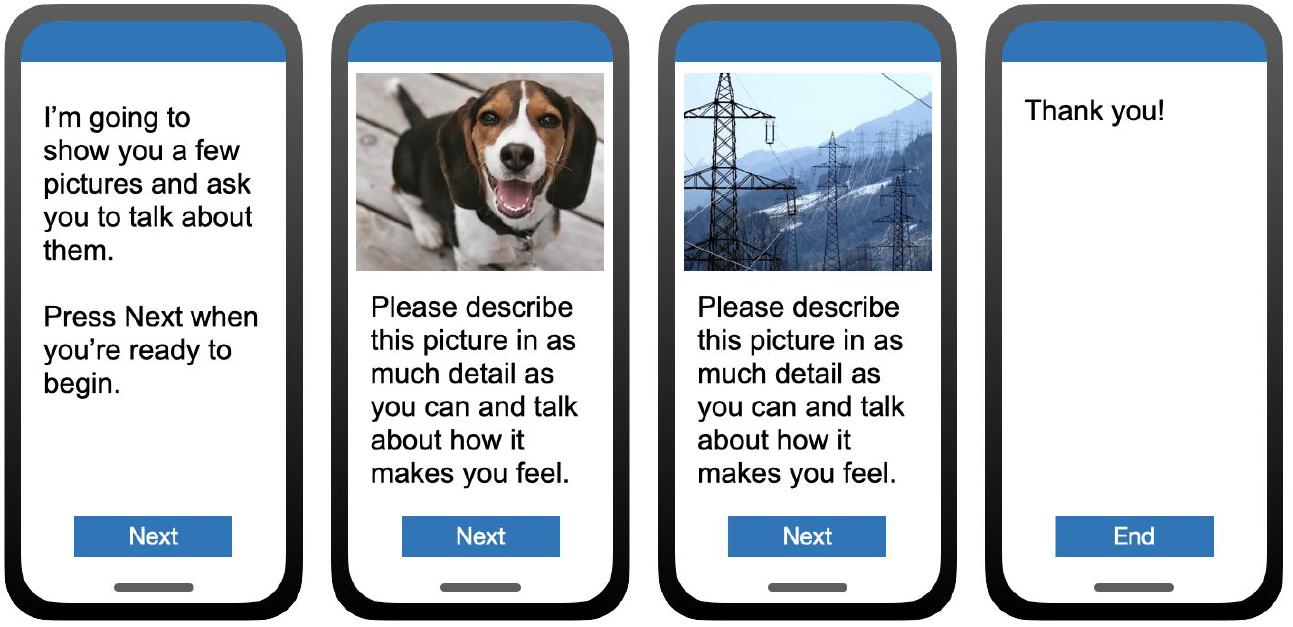
Depiction of the smartphone-based assessment that all individuals completed. Video and audio of participant responses was recorded during the assessment and used to quantify behavioral characteristics and subsequently measure digital markers of MDD severity.

### 2.4 Digital marker calculation

Video and audio were captured continuously during the smartphone assessment using the smartphone front-facing camera and microphone. Data was uploaded and processed through HIPAA compliant backend services for transfer and storage of Protected Health Information (PHI). Video was extracted for analysis for the portion of the task where the participant is observing the image and responding to it. Both video and audio was extracted and analysed for the portion of the task when the participant was describing the image.

All analysis was conducted in a Python environment using open-source tools. All code used to calculate visual and auditory markers of MDD, the resulting data output, and the executable python script for the analysis of the data output presented in this manuscript is publicly available on Github: https://github.com/AiCure/ms_dbm_adamsclinicalstudy. A total of 18 variables inclusive of the MADRS, facial markers, voice markers, and movement markers were calculated (see Supplementary Table 1 for descriptive statistics for all variables).

#### 2.4.1 Facial marker calculation

First, all videos were segmented into individual video frames at 30 frames per second. Next, each frame was segmented into three matrices consisting of red, blue, green spectrum pixels for use in computer vision (CV) modeling using OpenCV, a open source computer vision software package (Brahmbhatt, 2013). Subsequently, each frame was analysed using OpenFace (Baltrusaitis et al., 2016), an open source software package that has demonstrated validity next to expert human ratings of Facial Action Coding System (FACS) (Ekman and Rosenberg, 2005), a standardized methodology to measure facial movements that reflect activity in the underlying human facial musculature used in the production of basic emotions (i.e., *happiness, fear, anger, surprise, sadness, disgust)*.

Specifically, for each frame OpenFace outputs **(1)** binary activation of each facial action unit (AU) utilized to calculate presence of facial emotions and **(2)** the degree of expressivity for that action unit utilized to calculate intensity of facial emotions. From AU measurements, emotion behavior was calculated including **(1)** the presence or absence of each emotion for each frame selected as the most probable based on the observed AU activation, termed ‘count’ and **(2)** the level of activation for each emotion and across all emotions, termed ‘intensity’. Following the calculation of these variables for each frame, a set of variables was calculated that represented the count of emotions expressed across across all frames divided by number of frames (*fear count, anger count, surprise count, sadness count, happy count, disgust count)* and the intensity of emotion averaged over all frames (fear intensity, anger intensity, surprise intensity, sadness intensity, surprise intensity, disgust intensity). Additionally, a composite score of overall facial intensity summed across all emotions was calculated *(overall facial expressivity)*.

#### 2.4.2 Voice marker calculation

Recordings were segmented into speech and non-speech parts using parselmouth, an open source software package that utilizes Praat software library (Weenink and Boersma, 2018) functions for vocal analysis (Jadoul et al., 2018). The ratio of speech to white space between words was calculated to represent the amount of time participants spoke compared to non-speech (*voice percentage*).

#### 2.4.3 Movement marker calculation

For each frame of video, head position and angle were acquired using OpenFace. The average framewise displacement of the head between frames *(head movement mean)* and its standard deviation *(head movement standard deviation)* were calculated as measures of head movement. The mean change in angle of the head(head *pose change mean)* was calculated as an additional measure of head movement.

### 2.5 Data Analysis

Change over time in MADRS and facial, voice, and movement variables (termed digital markers) was calculated using repeated measures analysis of variance (ANOVA). To avoid capitalizing on change, p-values were corrected using false discovery rate (FDR) correction (Li and Barber, 2019). The sphericity assumption was formally tested for each ANOVA. When this assumption proved to hold, the F-statistic and corresponding p-value was used. When the sphericity assumption was violated, Mauchly’s W statistic and corresponding p-value were used (Mendoza, 1980). Additionally, pairwise comparisons were calculated between each time point to determine where change across time points occurs (i.e., baseline to 2 weeks, baseline to 4 weeks, 2 weeks to 4 weeks) controlling for FDR using Tukey’s test.

## 3 Results

### 3.1 Depression Response

Participants demonstrated a main effect for change in MADRS scores from baseline to week 4 [F(2,34) = 51.62, *p* < .0001]. Descriptive statistics demonstrate clinically relevant change with patients moving from the clinical to non-clinical range (Supplementary Table 1; Figure 2).

**Figure 2:**
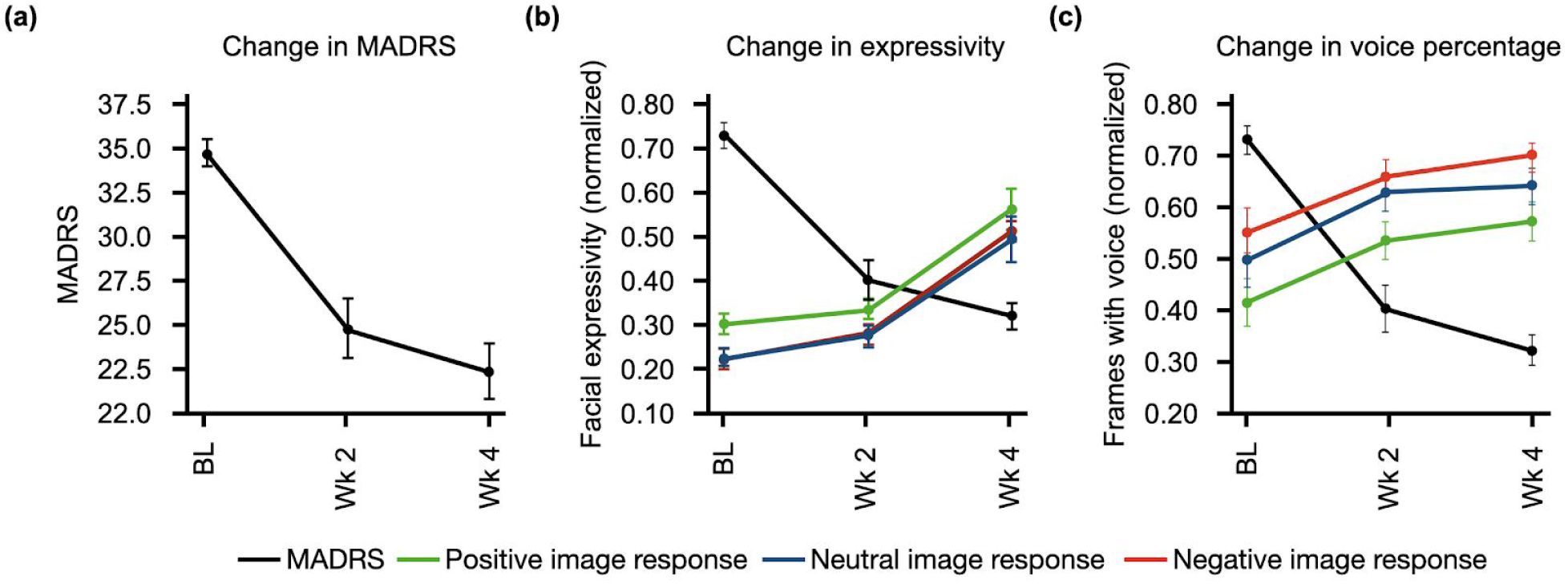
Response to treatment as measured by two independent assessments. Mean for each timepoint and standard error bars are shown. **(a)** MADRS scores acquired at baseline (BL), week 2, and week 4 showed significant decrease in response to ADT [F(2,34) = 51.62, p < .0001]. **(b)** Overall facial expressivity measured in response to positive, neutral, and negative images demonstrated a significant increase in response to ADT [F(2,28) = 39.27, p < .0001] as MADRS scores decreased. **(c)** Percentage of frames with voice measured in response to positive, neutral, and negative images also demonstrated a significant increase in response to ADT [F(2,26) = 5.77, p= 0.008] as MADRS scores decreased. All values in **b** and **c** were normalized between 0 and 1.

Participants demonstrated change in MDD severity as measured by digital markers. To align time points between digital markers and the MADRS scores, measurements from days 7-21 were averaged as the week 2 time point and measurements from days 22-35 were averaged as the week 4 time point. Due to missed remote assessments, a subset of the total sample of 18 had complete data across time points, with *n* = 12 for facial markers and *n* = 11 for voice markers. All statistical results for digital markers are presented in Table 1. Examples of marker profiles across treatment are presented in Figure 2 alongside the participants’ MADRS profile across treatment. All scores, including MADRS, were normalized to a range of 0-1 to allow visual comparison of the magnitude of change on digital markers in comparison to change in MADRS clinical scores (Figure 2).

**Table 1:** Repeated Measures ANOVA results for all facial and voice markers measured in response to positive, negative, and neutral visual stimuli from baseline to 4 weeks of ADT. Sph. indicates sphericity, F indicates F-statistic, P indicates p-value of F-statistic, and +/- indicates increased or decreased values.

|  | Neutral Stimuli |  |  |  | Positive Stimuli |  |  |  | Negative Stimuli |  |  |  |
| --- | --- | --- | --- | --- | --- | --- | --- | --- | --- | --- | --- | --- |
|  | Sph. | F | P | +/- | Sph. | F/W | P | +/- | Sph. | F | P | +/- |
| Voice percentage | T | 5.60 | .0095 | + | T | 3.59 | .0042 | + | T | 4.66 | .0187 | + |
| Anger intensity | T | 13.28 | <.0001 | + | T | 21.19 | <.0001 | + | T | 19.96 | <.0001 | + |
| Anger count | F | 2.54 | 0.1214 | n/a | F | .92 | 0.3787 | n/a | F | 2.40 | .1307 | n/a |
| Disgust intensity | T | 12.00 | .0002 | + | T | 9.00 | .0009 | + | T | 9.43 | .0007 | + |
| Disgust count | T | 0.51 | .6033 | n/a | T | 1.33 | .2796 | n/a | T | 0.31 | .7358 | n/a |
| Fear intensity | T | 41.23 | <.0001 | + | T | 32.50 | <.0001 | + | T | 60.38 | <.0001 | + |
| Fear count | F | 4.84 | .0413 | - | F | .67 | .5182 | n/a | F | .77 | .4287 | n/a |
| Happiness intensity | F | 6.03 | .0232 | + | F | 5.21 | .0306 | + | F | 4.38 | .0445 | + |
| Happiness count | T | .03 | .9666 | n/a | T | 0.46 | .6362 | n/a | F | 1.72 | .2089 | n/a |
| Sadness intensity | T | 13.53 | <.0001 | + | T | 8.67 | .0012 | + | T | 10.54 | .0004 | + |
| Sadness count | F | .59 | .4690 | n/a | T | 2.05 | .1473 | n/a | T | 1.90 | .16815 | n/a |
| Surprise intensity | T | 22.29 | <.0001 | + | T | 17.31 | <.0001 | + | T | 26.10 | <.0001 | + |
| Surprise count | T | 0.14 | .8665 | n/a | T | 0.20 | .8194 | n/a | T | 0.16 | .8497 | n/a |
| Overall expressivity | T | 32.60 | <.0001 | + | T | 40.67 | <.0001 | + | T | 36.95 | <.0001 | + |
| Head movement mean | F | 8.90 | .0069 | + | F | 3.58 | .0413 | + | F | 2.49 | .1335 | n/a |
| Head movement standard deviation | F | 3.68 | .0378 | + | T | 1.53 | .2333 | n/a | F | 1.59 | .2274 | n/a |
| Head pose change mean | F | 5.01 | .0325 | + | T | 3.18 | .0570 | + | F | 1.41 | .2595 | n/a |

### 3.2 Facial markers

All facial activity measures across all emotions (fear intensity, anger intensity, surprise intensity, sadness intensity, surprise intensity, disgust intensity and overall expressivity) along with the overall expressivity score demonstrated significant positive change from baseline to week 4 in response to all image prompts (positive, neutral, and negative; see Table 1). This result indicates that ADT produces a main effect on facial activity overall which is not bound to one particular facial musculature group or type of external stimulus (Figure 2).

Across conditions, the frequency of expressions of anger (anger count) decreases. The frequency of expressions of fear also decreases, but only in response to neutral and negative stimuli (fear count). Additionally, the frequency of expressions of happiness (happy count) decreases in response to negative stimuli only. Together, results indicate a general decrease in expressions of anger, and context specific decreases in fear and happiness expressions.

### 3.3 Voice markers

The single variable representing the ratio of speech to silence across sentences uttered (voice percentage) additionally demonstrated significant positive change in response to ADT across all conditions, indicating an increase in speech relative to silence. This result is consistent with increased motor/muscle activity observed in facial activity (Figure 2).

### 3.4 Movement markers

Additionally, movement parameters demonstrated consistent effects across conditions. The rate of head movement *(head movement mean)* and the degree of variability in the rate of head movement *(head movement standard deviation)* both demonstrated significant increases in response to ADT. *Head pose change mean* also demonstrated significant increase during neutral and positive stimuli.

## 4 Discussion

Results demonstrate a consistent effect of monoamine ADTs (SSRIs/SNRIs) on digital markers of motor functioning, which are highly concordant with change in MDD symptom severity. Specifically, facial and vocal activity demonstrated robust increases across 4 weeks following the initiation of treatment, which mirrored decreases in symptom severity as assessed by the clinician administered MADRS. The current findings suggest that SSRI/SNRI treatment, which produces graded increases in serotonin, reduces depression severity in part by rescuing motor functioning (e.g., increased facial expressivity, increased speech production).

Additionally, a decrease was observed across conditions in the expression of anger. Patients with depression have long demonstrated increased rates of anger compared to healthy counterparts (Koh et al., 2005; Riley et al., 1989). Further, polymorphisms of the Serotonin 1B receptor that are associated with increased depression and suicide risk are also associated with increased anger and fear (Hakulinen et al., 2013; McDevitt et al., 2011). These results further indicate that the observed change in digital markers in response to serotonin reuptake inhibitors reflects a more specific phenotypic change in measurement of serotonergic profile in the central nervous system.

Serotonin levels in the central nervous system are known to have both direct and indirect effects (via dopamine) on motor activity (Gerson and Baldessarini, 1980; Jacobs, 1991). Both suicide (as measured in post-mortem brain tissue) and suicidal attempts, a key symptom class of MDD, are associated with depleted serotonin (Mann and John Mann, 2013). As such, digital measurements that reflect motor behavior may represent a sensitive measure of serotonergic tone and potentially other neurotransmitter activity that affects motor functioning and ultimately the overall clinical presentation.

The current work presents a number of limitations that should be overcome through research that confirms and extends the findings reported. First, while treatment success was confirmed with clinical measures of MDD, dosage and treatment type were not controlled in a manner to make direct inferences about dose-response relationships. In addition, the current study was not adequately powered to assess the intra-subject variability in treatment response. Future research should provide more extensive experimental control of medication and dosage, and to assess the relationship between magnitude of clinical response and digital markers of motor activity. Next, while facial movement results were robust, we do not know if findings related to specific emotions would rise to significance given a larger sample size or more sampling occasions of the stimuli. One of the goals of the data collection was to implement a very simple remote assessment of objective visual and auditory markers to facilitate ease of frequent assessment. However, the minimum sample to accurately measure each marker needs to be assessed through the use of larger samples. For example, we observed decreases in happiness in response to negative images. This result is difficult to directly interpret. However, given a larger sample, we may be powered to identify increases in happiness in response to positive images, consistent with observations that depressed patients display context inappropriate affect (Coifman et al., 2007; Coifman and Bonanno, 2010).

Ultimately, the current work holds promise as an example of the potential to observe treatment effects that reflect underlying neurobiological target engagement by shifting the focus to monotonic neurobiologically based domains rather than heterogeneous diagnoses (Insel, 2014). Further work should determine if these same markers are relevant in other disorders and treatments that are mechanistically affected by serotonergic tone, as well as their relevance to other disorders with motor and movement profiles including Parkinson’s Disease and schizophrenia (Stahl, 2016). Second, the current work demonstrates the success of non-invasive objective digital assessment as a tool to assess treatment effects in Major Depressive Disorder. Importantly, no markers were scientifically novel, rather they were based on validated methods that are open and public and have been previously reported in scientific literature.

The current work demonstrates, in the context of MDD, that these data sources can be captured remotely through ubiquitously available digital tools to provide measurements that are at least as robust as traditional rating scales. It will be important to determine if such models reliably track with other disease states and treatment responses as such models and applications have significant potential to increase the rate and accuracy of treatment decision making.

Together, the current study demonstrates that scalability, through digital measurement, of monotonic characteristics that reflect underlying central nervous system activity. This observation holds promise that frequent remote digital assessment can be used to monitor, titrate, and even personalize treatment for MDD and other psychiatric or neurological conditions by grounding the measurements in narrow phenotypes that match the underlying mechanistic target of the treatment.

## Data Availability

All code used to calculate visual and auditory markers of MDD, the resulting data output, and the executable python script for the analysis of the data output presented in this manuscript is publicly available on Github.

https://github.com/AiCure/ms_dbm_adamsclinicalstudy

## Supplementary Materials

**Supplementary Table 1:** Means and standard errors across time points of each digital marker in response to neutral, positive and negative stimuli.

|  | Neutral Stimuli |  |  |  |  |  | Positive Stimuli |  |  |  |  |  | Negative Stimuli |  |  |  |  |  |
| --- | --- | --- | --- | --- | --- | --- | --- | --- | --- | --- | --- | --- | --- | --- | --- | --- | --- | --- |
| | $\mu$<br>W1 | S.E.<br>W1 | $\mu$<br>W2 | S.E.<br>W2 | $\mu$<br>W3 | S.E.<br>W3 | $\mu$<br>W1 | S.E.<br>W1 | $\mu$<br>W2 | S.E.<br>W2 | $\mu$<br>W3 | S.E.<br>W3 | $\mu$<br>W1 | S.E.<br>W1 | $\mu$<br>W2 | S.E.<br>W2 | $\mu$<br>W3 | S.E.<br>W3 |
| Voice percentage | 0.49 | 0.04 | 0.60 | 0.03 | 0.64 | 0.03 | 0.41 | 0.03 | 0.52 | 0.04 | 0.57 | 0.04 | 0.54 | 0.03 | 0.64 | 0.03 | 0.70 | 0.02 |
| Anger intensity | 0.28 | 0.01 | 0.30 | 0.02 | 0.46 | 0.04 | 0.30 | 0.02 | 0.33 | 0.02 | 0.47 | 0.03 | 0.34 | 0.01 | 0.37 | 0.02 | 0.54 | 0.04 |
| Anger count | 0.08 | 0.02 | 0.15 | 0.05 | 0.03 | 0.01 | 0.09 | 0.04 | 0.13 | 0.04 | 0.05 | 0.02 | 0.05 | 0.02 | 0.13 | 0.05 | 0.04 | 0.02 |
| Disgust intensity | 0.28 | 0.02 | 0.26 | 0.02 | 0.40 | 0.05 | 0.29 | 0.02 | 0.25 | 0.02 | 0.40 | 0.05 | 0.27 | 0.02 | 0.26 | 0.02 | 0.39 | 0.05 |
| Disgust count | 0.28 | 0.04 | 0.26 | 0.03 | 0.28 | 0.06 | 0.24 | 0.04 | 0.16 | 0.03 | 0.24 | 0.06 | 0.20 | 0.04 | 0.21 | 0.03 | 0.18 | 0.05 |
| Fear intensity | 0.21 | 0.02 | 0.26 | 0.02 | 0.55 | 0.05 | 0.24 | 0.02 | 0.27 | 0.02 | 0.50 | 0.04 | 0.17 | 0.01 | 0.21 | 0.02 | 0.48 | 0.05 |
| Fear count | 0.10 | 0.04 | 0.05 | 0.01 | 0.03 | 0.03 | 0.04 | 0.03 | 0.01 | 0.003 | 0.02 | 0.01 | 0.10 | 0.04 | 0.04 | 0.02 | 0.02 | 0.01 |
| Happiness intensity | 0.12 | 0.02 | 0.13 | 0.03 | 0.26 | 0.06 | 0.15 | 0.03 | 0.12 | 0.03 | 0.27 | 0.07 | 0.13 | 0.03 | 0.12 | 0.03 | 0.25 | 0.07 |
| Happiness count | 0.07 | 0.02 | 0.10 | 0.04 | 0.09 | 0.03 | 0.06 | 0.02 | 0.10 | 0.03 | 0.09 | 0.04 | 0.10 | 0.03 | 0.11 | 0.04 | 0.10 | 0.05 |
| Sadness intensity | 0.20 | 0.02 | 0.23 | 0.02 | 0.40 | 0.05 | 0.30 | 0.02 | 0.32 | 0.02 | 0.47 | 0.04 | 0.21 | 0.02 | 0.24 | 0.02 | 0.38 | 0.05 |
| Sad count | 0.12 | 0.03 | 0.08 | 0.01 | 0.12 | 0.06 | 0.07 | 0.02 | 0.06 | 0.02 | 0.08 | 0.03 | 0.19 | 0.04 | 0.13 | 0.03 | 0.16 | 0.05 |
| Surprise intensity | 0.41 | 0.03 | 0.42 | 0.05 | 0.69 | 0.04 | 0.42 | 0.03 | 0.42 | 0.04 | 0.65 | 0.04 | 0.39 | 0.03 | 0.42 | 0.05 | 0.66 | 0.04 |
| Surprise count | 0.30 | 0.04 | 0.24 | 0.03 | 0.29 | 0.06 | 0.26 | 0.05 | 0.19 | 0.03 | 0.23 | 0.04 | 0.27 | 0.04 | 0.21 | 0.03 | 0.19 | 0.03 |
| Head velocity | 0.22 | 0.03 | 0.21 | 0.03 | 0.26 | 0.04 | 0.21 | 0.03 | 0.23 | 0.04 | 0.23 | 0.04 | 0.16 | 0.03 | 0.18 | 0.03 | 0.21 | 0.04 |
| Standard deviation of head velocity | 0.26 | 0.04 | 0.20 | 0.03 | 0.29 | 0.06 | 0.26 | 0.04 | 0.23 | 0.03 | 0.27 | 0.05 | 0.25 | 0.04 | 0.21 | 0.04 | 0.27 | 0.07 |
| Head pose | 0.25 | 0.04 | 0.22 | 0.03 | 0.30 | 0.06 | 0.11 | 0.03 | 0.08 | 0.01 | 0.09 | 0.02 | 0.20 | 0.03 | 0.20 | 0.03 | 0.22 | 0.06 |
| Overall expressivity | 0.22 | 0.02 | 0.26 | 0.02 | 0.49 | 0.05 | 0.30 | 0.02 | 0.31 | 0.02 | 0.56 | 0.05 | 0.23 | 0.02 | 0.26 | 0.03 | 0.51 | 0.05 |

*Note:* $\mu$ = mean score; W= week of trial; S.E.= standard error of the estimate

**Supplementary Table 2:** Pairwise comparisons between time points for each digital marker in response to neutral, positive and negative stimuli.

| Variable | Neutral stimuli |  |  |  | Positive stimuli |  |  |  | Negative Stimuli |  |  |  |
| --- | --- | --- | --- | --- | --- | --- | --- | --- | --- | --- | --- | --- |
|  | margin | SE | Tukey |  | margin | SE | Tukey |  | margin | SE | Tukey |  |
|  |  |  | T | p |  |  | T | p |  |  | T | p |
| Voice Percentage |  |  |  |  |  |  |  |  |  |  |  |  |
| - Week 1 and Week 3 | -0.120 | 0.068 | -1.763 | 0.185 | -0.137 | 0.071 | -1.945 | 0.129 | -0.106 | 0.059 | -1.793 | 0.175 |
| - Week 1 and Week 5 | -0.123 | 0.071 | -1.730 | 0.197 | -0.139 | 0.074 | -1.879 | 0.148 | -0.131 | 0.062 | -2.095 | 0.094 |
| - Week 3 and Week 5 | -0.003 | 0.072 | -0.044 | 0.900 | -0.001 | 0.075 | -0.019 | 0.900 | -0.024 | 0.063 | -0.375 | 0.900 |
| Anger intensity |  |  |  |  |  |  |  |  |  |  |  |  |
| - Week 1 and Week 3 | -0.052 | 0.043 | -1.196 | 0.458 | -0.072 | 0.044 | -1.625 | 0.237 | -0.052 | 0.046 | -1.132 | 0.495 |
| - Week 1 and Week 5 | -0.219 | 0.045 | -4.852 | 0.001 | -0.217 | 0.046 | -4.714 | 0.001 | -0.240 | 0.047 | -5.055 | 0.001 |
| - Week 3 and Week 5 | -0.167 | 0.046 | -3.647 | 0.001 | -0.145 | 0.047 | -3.106 | 0.006 | -0.188 | 0.048 | -3.907 | 0.001 |
| Anger count |  |  |  |  |  |  |  |  |  |  |  |  |
| - Week 1 and Week 3 | -0.049 | 0.046 | -1.051 | 0.542 | -0.050 | 0.050 | -1.007 | 0.568 | -0.078 | 0.049 | -1.588 | 0.253 |
| - Week 1 and Week 5 | 0.028 | 0.048 | 0.581 | 0.813 | -0.004 | 0.052 | -0.086 | 0.900 | -0.034 | 0.051 | -0.656 | 0.770 |
| - Week 3 and Week 5 | 0.077 | 0.049 | 1.565 | 0.263 | 0.046 | 0.053 | 0.866 | 0.649 | 0.045 | 0.052 | 0.854 | 0.656 |
| Disgust intensity |  |  |  |  |  |  |  |  |  |  |  |  |
| - Week 1 and Week 3 | 0.033 | 0.054 | 0.611 | 0.796 | 0.045 | 0.055 | 0.822 | 0.674 | -0.032 | 0.054 | -0.595 | 0.805 |
| - Week 1 and Week 5 | -0.117 | 0.057 | -2.056 | 0.102 | -0.115 | 0.057 | -2.014 | 0.111 | -0.141 | 0.056 | -2.518 | 0.033 |
| - Week 3 and Week 5 | -0.150 | 0.058 | -2.601 | 0.027 | -0.160 | 0.058 | -2.760 | 0.017 | -0.109 | 0.057 | -1.917 | 0.137 |
| Disgust count |  |  |  |  |  |  |  |  |  |  |  |  |
| - Week 1 and Week 3 | -0.005 | 0.079 | -0.062 | 0.900 | 0.110 | 0.083 | 1.313 | 0.390 | -0.056 | 0.074 | -0.759 | 0.711 |
| - Week 1 and Week 5 | -0.023 | 0.082 | -0.276 | 0.900 | 0.033 | 0.087 | 0.375 | 0.900 | -0.031 | 0.077 | -0.404 | 0.900 |
| - Week 3 and Week 5 | -0.018 | 0.083 | -0.213 | 0.900 | -0.077 | 0.088 | -0.871 | 0.646 | 0.025 | 0.079 | 0.320 | 0.900 |
| Fear intensity |  |  |  |  |  |  |  |  |  |  |  |  |
| - Week 1 and Week 3 | -0.076 | 0.051 | -1.491 | 0.298 | -0.070 | 0.049 | -1.411 | 0.337 | -0.069 | 0.048 | -1.438 | 0.323 |
| - Week 1 and Week 5 | -0.397 | 0.053 | -7.477 | 0.001 | -0.323 | 0.051 | -6.281 | 0.001 | -0.365 | 0.050 | -7.266 | 0.001 |
| - Week 3 and Week 5 | -0.321 | 0.054 | -5.953 | 0.001 | -0.254 | 0.052 | -4.851 | 0.001 | -0.296 | 0.051 | -5.795 | 0.001 |
| Fear count |  |  |  |  |  |  |  |  |  |  |  |  |
| - Week 1 and Week 3 | 0.100 | 0.062 | 1.621 | 0.239 | 0.015 | 0.016 | 0.972 | 0.588 | 0.019 | 0.056 | 0.339 | 0.900 |
| - Week 1 and Week 5 | 0.134 | 0.064 | 2.086 | 0.095 | 0.002 | 0.016 | 0.094 | 0.900 | 0.063 | 0.058 | 1.096 | 0.516 |
| - Week 3 and Week 5 | 0.034 | 0.065 | 0.523 | 0.847 | -0.014 | 0.017 | -0.825 | 0.673 | 0.045 | 0.059 | 0.759 | 0.711 |
| Happiness intensity |  |  |  |  |  |  |  |  |  |  |  |  |
| - Week 1 and Week 3 | -0.002 | 0.057 | -0.040 | 0.900 | 0.017 | 0.068 | 0.253 | 0.900 | -0.005 | 0.058 | -0.080 | 0.900 |
| - Week 1 and Week 5 | -0.116 | 0.059 | -1.966 | 0.123 | -0.118 | 0.070 | -1.674 | 0.218 | -0.112 | 0.060 | -1.868 | 0.151 |
| - Week 3 and Week 5 | -0.114 | 0.060 | -1.898 | 0.142 | -0.135 | 0.072 | -1.887 | 0.145 | -0.107 | 0.061 | -1.763 | 0.185 |
| Happiness count |  |  |  |  |  |  |  |  |  |  |  |  |
| - Week 1 and Week 3 | -0.004 | 0.036 | -0.112 | 0.9 | -0.019 | 0.043 | -0.449 | 0.889 | 0.011 | 0.054 | 0.202 | 0.900 |
| - Week 1 and Week 5 | -0.004 | 0.037 | -0.097 | 0.9 | 0.002 | 0.045 | 0.056 | 0.900 | 0.042 | 0.057 | 0.745 | 0.719 |
| - Week 3 and Week 5 | 0.000 | 0.038 | 0.010 | 0.9 | 0.022 | 0.045 | 0.479 | 0.872 | 0.031 | 0.058 | 0.543 | 0.835 |
| Sadness intensity |  |  |  |  |  |  |  |  |  |  |  |  |
| - Week 1 and Week 3 | -0.020 | 0.049 | -0.414 | 0.900 | 0.012 | 0.043 | 0.270 | 0.900 | -0.065 | 0.050 | -1.309 | 0.392 |
| - Week 1 and Week 5 | -0.191 | 0.051 | -3.749 | 0.001 | -0.137 | 0.045 | -3.040 | 0.007 | -0.194 | 0.052 | -3.717 | 0.001 |
| - Week 3 and Week 5 | -0.171 | 0.052 | -3.301 | 0.003 | -0.148 | 0.046 | -3.247 | 0.004 | -0.128 | 0.053 | -2.423 | 0.043 |
| Sadness count |  |  |  |  |  |  |  |  |  |  |  |  |
| - Week 1 and Week 3 | 0.011 | 0.066 | 0.166 | 0.900 | -0.046 | 0.041 | -1.115 | 0.505 | -0.004 | 0.078 | -0.050 | 0.900 |
| - Week 1 and Week 5 | -0.052 | 0.069 | -0.762 | 0.709 | -0.078 | 0.043 | -1.840 | 0.160 | -0.089 | 0.081 | -1.100 | 0.514 |
| - Week 3 and Week 5 | -0.063 | 0.070 | -0.907 | 0.625 | -0.033 | 0.043 | -0.758 | 0.711 | -0.085 | 0.082 | -1.036 | 0.551 |
| Surprise intensity |  |  |  |  |  |  |  |  |  |  |  |  |
| - Week 1 and Week 3 | -0.030 | 0.077 | -0.394 | 0.900 | -0.061 | 0.069 | -0.891 | 0.635 | -0.054 | 0.078 | -0.691 | 0.750 |
| - Week 1 and Week 5 | -0.312 | 0.080 | -3.898 | 0.001 | -0.288 | 0.072 | -4.007 | 0.001 | -0.292 | 0.082 | -3.584 | 0.001 |
| - Week 3 and Week 5 | -0.282 | 0.081 | -3.465 | 0.002 | -0.227 | 0.073 | -3.104 | 0.006 | -0.238 | 0.083 | -2.876 | 0.012 |
| Surprise count |  |  |  |  |  |  |  |  |  |  |  |  |
| - Week 1 and Week 3 | -0.001 | 0.073 | -0.018 | 0.9 | 0.004 | 0.071 | 0.056 | 0.900 | -0.023 | 0.064 | -0.361 | 0.900 |
| - Week 1 and Week 5 | -0.027 | 0.076 | -0.350 | 0.9 | -0.029 | 0.074 | -0.398 | 0.900 | 0.005 | 0.067 | 0.075 | 0.900 |
| - Week 3 and Week 5 | -0.025 | 0.077 | -0.327 | 0.9 | -0.033 | 0.075 | -0.445 | 0.892 | 0.028 | 0.068 | 0.415 | 0.900 |
| Overall expressivity |  |  |  |  |  |  |  |  |  |  |  |  |
| - Week 1 and Week 3 | -0.037 | 0.052 | -0.722 | 0.732 | -0.032 | 0.053 | -0.609 | 0.797 | -0.059 | 0.054 | -1.096 | 0.516 |
| - Week 1 and Week 5 | -0.284 | 0.054 | -5.259 | 0.001 | -0.288 | 0.055 | -5.213 | 0.001 | -0.305 | 0.056 | -5.454 | 0.001 |
| - Week 3 and Week 5 | -0.246 | 0.055 | -4.495 | 0.001 | -0.256 | 0.056 | -4.557 | 0.001 | -0.246 | 0.057 | -4.334 | 0.001 |
| Head velocity |  |  |  |  |  |  |  |  |  |  |  |  |
| - Week 1 and Week 3 | 0.008 | 0.049 | 0.164 | 0.900 | 0.016 | 0.049 | 0.333 | 0.900 | -0.026 | 0.053 | -0.481 | 0.871 |
| - Week 1 and Week 5 | -0.099 | 0.051 | -1.950 | 0.128 | -0.053 | 0.051 | -1.039 | 0.549 | -0.072 | 0.055 | -1.298 | 0.398 |
| - Week 3 and Week 5 | -0.107 | 0.052 | -2.074 | 0.098 | -0.070 | 0.052 | -1.337 | 0.377 | -0.046 | 0.056 | -0.824 | 0.673 |
| Standard deviation of head velocity |  |  |  |  |  |  |  |  |  |  |  |  |
| - Week 1 and Week 3 | 0.021 | 0.073 | 0.287 | 0.900 | 0.030 | 0.071 | 0.429 | 0.900 | -0.028 | 0.094 | -0.294 | 0.900 |
| - Week 1 and Week 5 | -0.110 | 0.076 | -1.449 | 0.318 | -0.047 | 0.074 | -0.638 | 0.780 | -0.120 | 0.098 | -1.228 | 0.439 |
| - Week 3 and Week 5 | -0.131 | 0.077 | -1.697 | 0.209 | -0.078 | 0.075 | -1.033 | 0.552 | -0.092 | 0.099 | -0.930 | 0.612 |
| Head pose |  |  |  |  |  |  |  |  |  |  |  |  |
| - Week 1 and Week 3 | -0.025 | 0.070 | -0.363 | 0.900 | 0.020 | 0.020 | 0.976 | 0.586 | -0.015 | 0.082 | -0.183 | 0.900 |
| - Week 1 and Week 5 | -0.142 | 0.073 | -1.949 | 0.128 | -0.012 | 0.021 | -0.546 | 0.833 | -0.094 | 0.085 | -1.098 | 0.515 |
| - Week 3 and Week 5 | -0.116 | 0.074 | -1.576 | 0.258 | -0.031 | 0.021 | -1.459 | 0.313 | -0.079 | 0.087 | -0.908 | 0.625 |

